# Changes in SARS-CoV-2 seroprevalence and population immunity in Finland, 2020–2022

**DOI:** 10.1101/2023.02.17.23286042

**Authors:** Anna Solastie, Tuomo Nieminen, Nina Ekström, Hanna Nohynek, Lasse Lehtonen, Arto A. Palmu, Merit Melin

## Abstract

Studying the prevalence of SARS-CoV-2 specific antibodies (seroprevalence) allows assessing the impact of epidemic containment measures and vaccinations, as well as estimation of the number of infections regardless of viral testing. We assessed antibody-mediated immunity to SARS-CoV-2 induced by infections and vaccinations from April 2020 to December 2022 in Finland by measuring serum IgG to SARS-CoV-2 nucleoprotein (N-IgG) and spike glycoprotein from randomly selected 18-85-year-old subjects (n=9794). N-IgG seroprevalence remained at <7% until the last quartile (Q) of 2021. After the emergence of the omicron variant, N-IgG seroprevalence increased rapidly and was 31% in Q1/2022 and 54% in Q4/2022. Seroprevalence was highest in youngest age groups from Q2/2022 onwards. We estimated that 51% of the Finnish 18-85-year-old population had antibody-mediated hybrid immunity induced by a combination of vaccinations and infections by the end of 2022. In conclusion, major shifts in the COVID-19 pandemic and resulting population immunity could be observed by serological testing.

## 2. Background

The severe acute respiratory syndrome coronavirus 2 (SARS-CoV-2), responsible for the coronavirus disease 2019 (COVID-19) pandemic^1^, has spread rapidly in successive waves and caused significant morbidity and mortality throughout the world^2^. However, the epidemic waves and different SARS-CoV-2 variants have progressed at varying pace and volume in different countries^3–5^.

Estimates of the proportion of individuals with antibodies to SARS-CoV-2 (seroprevalence) can provide important, timely information on the dynamics of exposure and immunity to the virus in the population over time. This is critical for evidence-based decision-making regarding containment measures and vaccination strategies. Continuous estimation of seroprevalence (serosurveillance) can also be used to assess the impact of containment measures, the effectiveness of vaccination programmes, and the effects of emerging variants on infections.

One way to estimate the incidence of SARS-CoV-2 infections is through the number of laboratory-confirmed infections. However, during the early stages of the COVID-19 epidemic in Finland, access to COVID-19 testing was very limited, and other methods were needed to estimate the incidence of the infection. In Finland, a state of emergency was declared on March 16, 2020 resulting in extensive restrictions in contacts. The Finnish Institute for Health and Welfare initiated a study^6^ to find out how widespread SARS-CoV-2 infections were in the population and how many had developed immunity against the virus. Samples were collected and analysed, and the results were initially reported weekly in 2020. Early results suggested that likely less than 1% of the population was infected before summer 2020 and the true numbers of infections until then were 1 to 5 times those diagnosed by virus testing^7^. Seroprevalence estimates served as indicators of the progress of the epidemic in Finland and guided public health decision-making.

In December 2020 mass COVID-19 vaccinations began in Finland, with healthcare workers, the elderly, and others at the highest risk of severe COVID-19 being the first to whom vaccines were offered. COVID-19 vaccinations were gradually extended so that by July 2021 all 18-year-olds and older were eligible. Although PCR testing capacity caught on and was widely accessible since August 2020 in Finland, it remained important to follow population immunity and estimate whether viral testing captured most infections. In late-2021 testing strategies changed to less comprehensive PCR testing^8,9^ due to the availability of the at-home antigen tests and the rapid increase in infections caused by the Omicron variant^10,11^. However, at-home antigen test results were not reported to any register, and serology-based estimations on number of SARS-CoV-2 infections became indispensable again.

Monitoring the gradual development of the population immunity elicited by SARS-CoV-2 infections and/or vaccinations has been crucial in the planning of epidemic containment measures and vaccination program in Finland. Here, we present the results of a continuing serosurveillance study spanning from April 2020 to December 2022, focusing on the effects of Omicron emergence on the Finnish adult population seroprevalence and hybrid immunity.

## 3. Results

### 3.1 Description of the study population

Of the 34 619 invited subjects, 9794 (28%) donated sera from April 2020 to December 2022. Participation decreased during the study from 64% in the first sample in 2020 to 19% in the last sample of 2022 (Table S1). The distribution of sample collection per month and year is presented in Table S2. Participant demographics and comparison to the adult population in Finland and the capital region (Uusimaa) are summarised in Table S3. More females (61%) than males (39%) donated sera. Compared to Finland’s nationwide age and native language distributions, 18-29-year-olds participated less frequently, whilst the 45-64-year-old and native Finnish or Swedish speaker groups were disproportionally large (Table S3). Additionally, 96% of the participants had received at least one COVID-19 vaccine before 2022, exceeding the 88% vaccination rate of 18-85-year-olds in Finland during the same period (Table S3). Most samples (54%) were collected in Finland’s capital region (Helsinki and Uusimaa healthcare district), but regional distributions varied with random population surveys (Table S1, Table S4).

### 3.2 Assay sensitivity in detection of past SARS-CoV-2 infection

For samples collected 0-5 months, 6-12 months and over 12 months after PCR-confirmed infection, 90% (n=291), 74% (n=146) and 65% (n=26) had N-IgG, respectively. From March 2022 onwards participants were asked if they had received a positive at-home antigen test result. When we combined the N-IgG results of those with a positive PCR and/or at-home antigen test, the estimates remained similar. The N-IgG assay sensitivity in the detection of past SARS-CoV-2 infection confirmed by either PCR or at-home antigen test was 91% at 0-5 months (n=641), 74% at 6-12 months (n=292) and 62% at over 12 months (n=21) after the most recent positive test. S-IgG assay sensitivity estimated from unvaccinated subjects was 82% (n=62) for samples taken up to 12 months after infection. S-IgG assay sensitivity could not be assessed for samples taken over a year after infection (n=2). S-IgG sensitivity in the detection of antibodies induced by vaccination alone or vaccination and infection was 99% (n=3288) including all samples from subjects with at least one dose ≥14 days before sample collection, the last of which was collected 559 days after the most recent COVID-19 vaccine dose. We were unable to assess the effect of possible reinfections as we did not have repeated samples from the subjects.

### 3.3 N-IgG seroprevalence trends in 2020-2022

During 2020, age-adjusted N-IgG seroprevalence among Finnish adults was 2% [95% confidence interval 1-4%] in April-June (Q2), 4% [2-6%] in July-September (Q3) and 2% [1-8%] in October-December (Q4) (Figure 1, Table 1). In 2021 the age-adjusted N-IgG seroprevalence of Finnish adults remained at under 7% (Table 1) and was highest among 30-44-year-olds in Q4/2021 (Figure 2, Table S5). In Q1/2022 we observed a steep increase in N-IgG seroprevalence (Figure 1); already 31% [19-45%] of Finnish adults had N-IgG (Table 1), with 30-44-year-olds having the highest seroprevalence (41%) (Figure 2, Table S5). During 2022 N-IgG seroprevalence continued to increase in all age groups (Figure 2). From Q2/2022 onwards N-IgG seroprevalence was the highest in 18-29-year-olds and decreased gradually with increasing age (Figure 2, Table S5). In Q4/2022 age-adjusted N-IgG seroprevalence was 54% [38-68%] for Finnish adults overall (Figure 1, Table 1) and 71% for the 18-29-year-olds (Table S5).

**Table 1.**
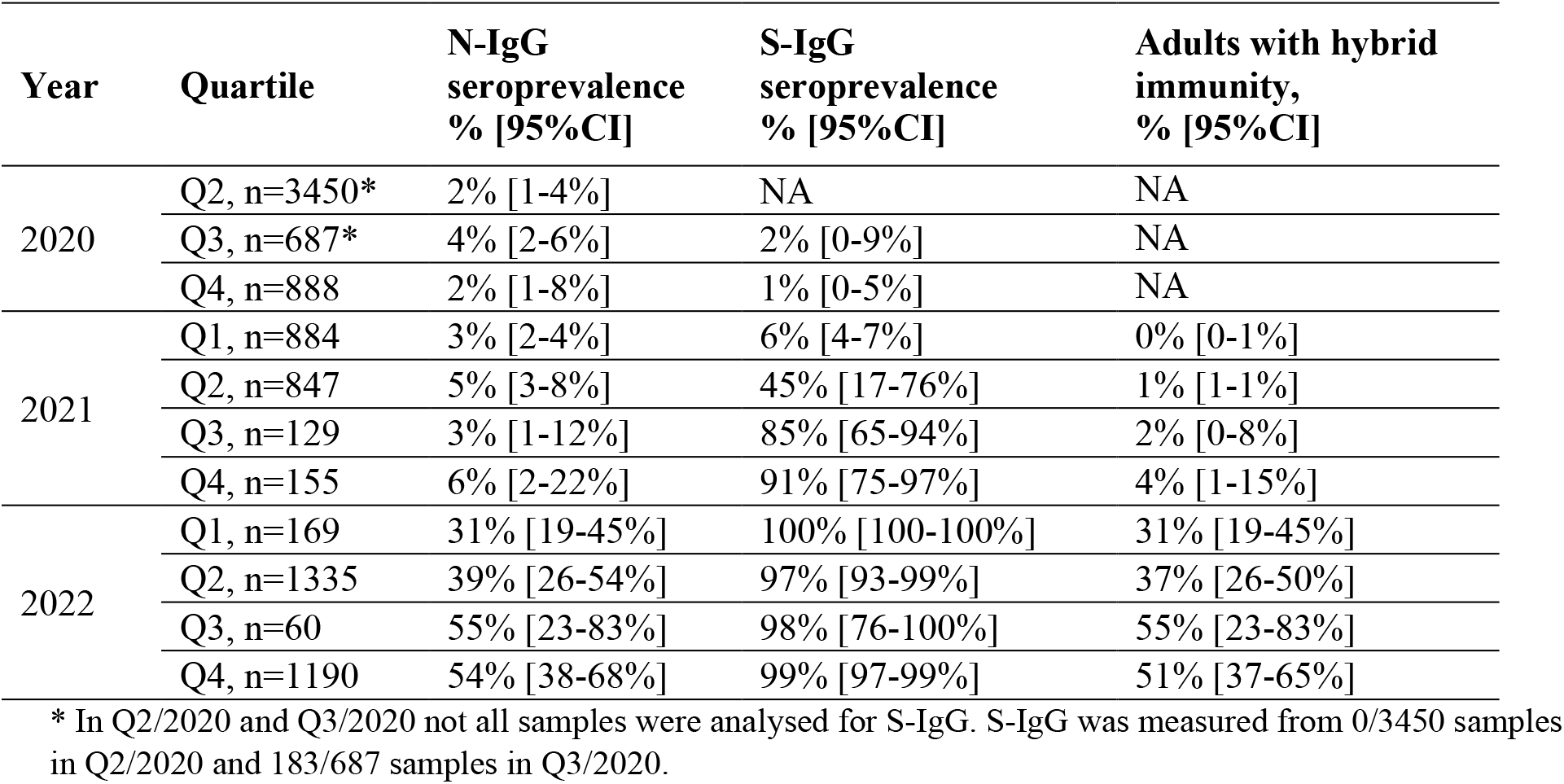
Age-adjusted N-IgG and S-IgG seroprevalence and antibody-mediated hybrid immunity in Finnish adults (18-85-year-olds) with 95% confidence intervals (CI).

| Year | Quartile | N-IgG seroprevalence % [95%CI] | S-IgG seroprevalence % [95%CI] | Adults with hybrid immunity, % [95%CI] |
| --- | --- | --- | --- | --- |
| 2020 | Q2, n=3450* | 2% [1-4%] | NA | NA |
|  | Q3, n=687* | 4% [2-6%] | 2% [0-9%] | NA |
|  | Q4, n=888 | 2% [1-8%] | 1% [0-5%] | NA |
| 2021 | Q1, n=884 | 3% [2-4%] | 6% [4-7%] | 0% [0-1%] |
|  | Q2, n=847 | 5% [3-8%] | 45% [17-76%] | 1% [1-1%] |
|  | Q3, n=129 | 3% [1-12%] | 85% [65-94%] | 2% [0-8%] |
|  | Q4, n=155 | 6% [2-22%] | 91% [75-97%] | 4% [1-15%] |
| 2022 | Q1, n=169 | 31% [19-45%] | 100% [100-100%] | 31% [19-45%] |
|  | Q2, n=1335 | 39% [26-54%] | 97% [93-99%] | 37% [26-50%] |
|  | Q3, n=60 | 55% [23-83%] | 98% [76-100%] | 55% [23-83%] |
|  | Q4, n=1190 | 54% [38-68%] | 99% [97-99%] | 51% [37-65%] |

\* In Q2/2020 and Q3/2020 not all samples were analysed for S-IgG. S-IgG was measured from 0/3450 samples in Q2/2020 and 183/687 samples in Q3/2020.

**Figure 1.**
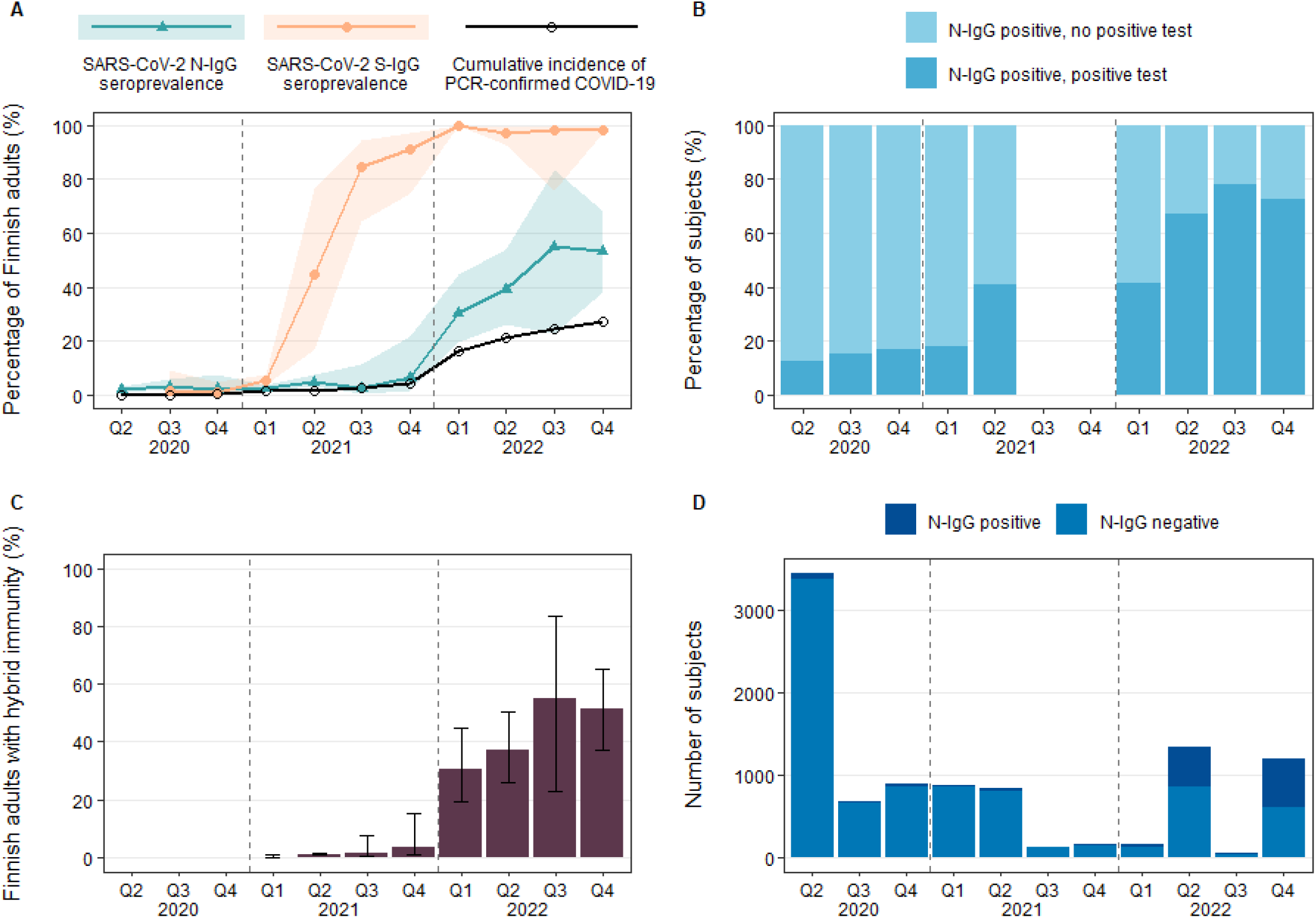
Population immunity in Finnish adults from April 2020 to December 2022 in quartiles (Q1-Q4). Dashed vertical lines divide calendar years. In 2020-2021 the oldest age group comprised of 65-70-year-olds and was extended to 65-85-year-olds in 2022. **A**. Age-adjusted SARS-CoV-2 nucleoprotein (N-IgG) and spike glycoprotein (S-IgG) seroprevalence and cumulative incidence of PCR-confirmed COVID-19 in Finnish adults. Error bars represent 95% confidence intervals. Samples collected before Q3/2020 were not analysed for S-IgG. **B**. N-IgG positive subjects divided into two groups; those without a positive PCR- or at-home antigen test (no positive test) and those with PCR- and/or at-home antigen test confirmed COVID-19 (positive test). Timepoints in which some age groups did not have N-IgG positives are not shown. **C**. Antibody-mediated hybrid immunity rates (%). Subjects with hybrid immunity had received at least one dose ≥14 days before sampling and were N-IgG and S-IgG positive. Error bars represent 95% confidence intervals. **D**. Number of subjects by N-IgG status.

**Figure 2.**
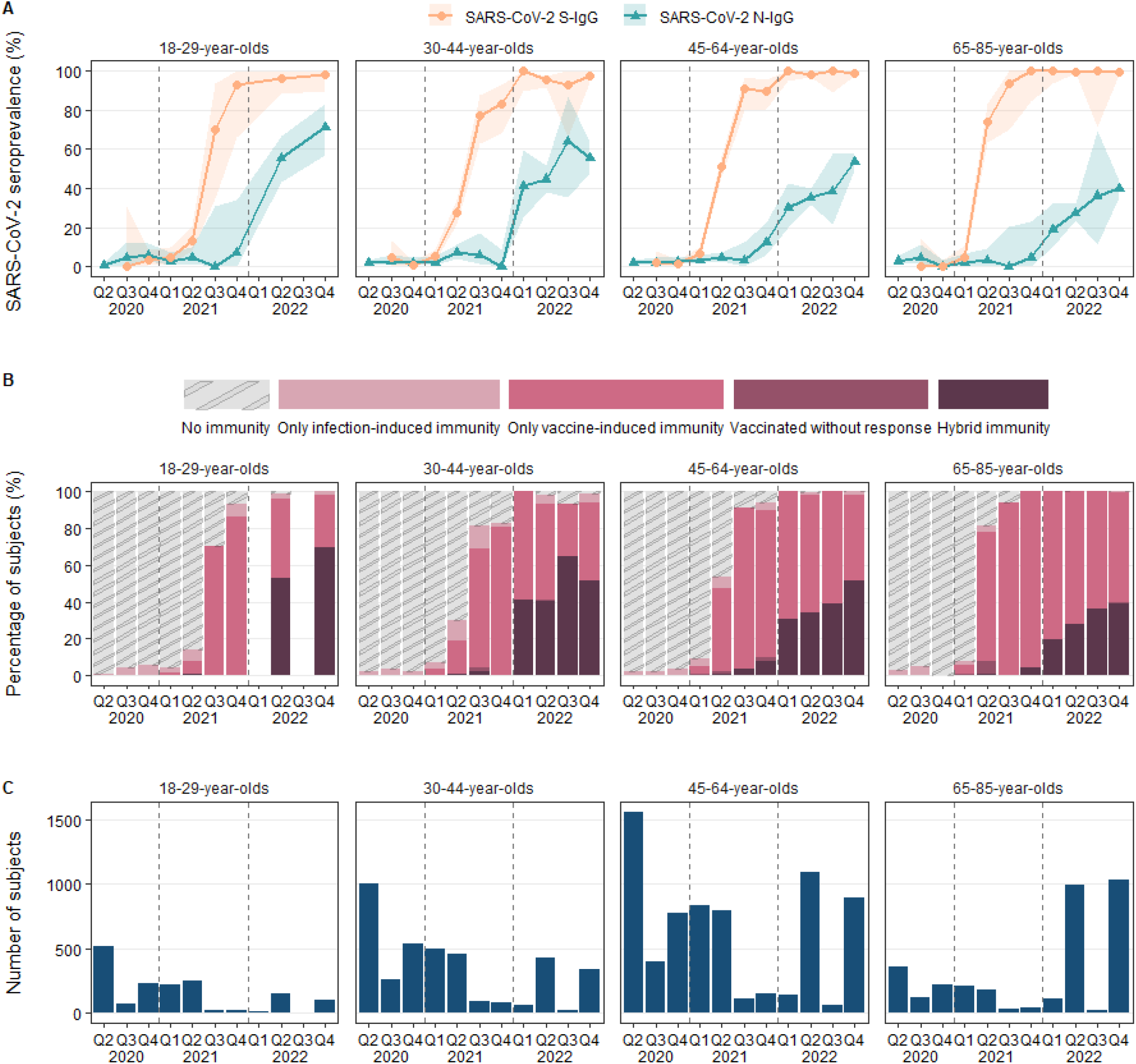
SARS-CoV-2 seroprevalence in Finnish adults from April 2020 to December 2022 in quartiles (Q1-Q4) by age groups. Dashed vertical lines divide calendar years. In 2020-2021 the oldest age group comprised of 65-70-year-olds and was extended to 65-85-year-olds in 2022. **A**. Development of SARS-CoV-2 nucleoprotein (N-IgG) and spike glycoprotein (S-IgG) seroprevalence in Finland by age group. Error bars represent 95% confidence intervals (Clopper-Pearson). Samples collected before Q3/2020 were not analysed for S-IgG. Timepoints with <10 samples per age group are not shown. **B**. Antibody-mediated of immunity by calendar year quartile. No immunity = N-IgG and S-IgG negative, unvaccinated before sampling. Only infection-induced immunity = Unvaccinated before sampling and N-IgG and/or S-IgG positive. Only vaccine-induced immunity = Had received at least one dose ≥14 days before sampling, S-IgG positive but N-IgG negative. Vaccinated without response = Had received at least one dose ≥14 days before sampling but were S-IgG negative. Hybrid immunity = Had received at least one dose ≥14 days before sampling and were N-IgG and S-IgG positive. Timepoints with <10 samples per age group are not shown. **C**. Number of samples in each age group and calendar year quartile.

### 3.4 S-IgG seroprevalence trends in 2020-2022

Age-adjusted S-IgG seroprevalence remained below 3% through 2020 (Figure 1, Table 1). In Q2/2021 13%, 28%, 51% and 74% of 18-29, 30-44, 45-64 and 65-85-year-old subjects, respectively, had S-IgG (Table 1, Figure 2). S-IgG seroprevalence increased steeply during 2021, reached 85% by Q4/2021, and stayed at over 90% until the end of follow-up in late 2022 (Figure 1, Table 1). In Q4/2022, S-IgG seroprevalence in Finnish adults was 99% [97-99%] (Table 1). Among the unvaccinated S-IgG seroprevalence was 2% [1-4%] in Q1/2021 and 9% [5-15%] in Q2/2021. From July 2021 onwards most participants had received at least one dose before sampling (n=2937/3038), and after mid-2021 S-IgG seroprevalence could not be reliably estimated for the unvaccinated.

### 3.5 Antibody-mediated immunity

In 2020 most subjects had no antibody-mediated immunity to COVID-19, and in 2021 most had only vaccine-induced immunity (Figure 2). Vaccine and infection-induced immunity (hybrid immunity) levels remained low until the beginning of 2022 when hybrid immunity increased from 4% to 31% between Q4/2021 and Q1/2022 Q1 (Figure 1, Table 1). In Q2 53% and in Q4/2022 71% of 18-29-year-olds had hybrid immunity (Figure 2). The lowest hybrid immunity proportions were observed for 65-85-year-olds, of whom 39% had hybrid immunity by the end of 2022 (Figure 2). We estimated that 37% of the Finnish adults had hybrid immunity in Q2/2022 and by Q4/2022 the proportion had increased to 51% (Figure 1, Table 1).

### 3.6 Test-confirmed infections in 2020-2022

In Q2/2020 under 15% of subjects who were N-IgG positive had a PCR-confirmed infection before sample collection (Figure 1). The proportion of subjects with test-confirmed infection increased slowly over time. By Q2/2021 41% of N-IgG positive subjects had known of their infection by PCR test (Figure 1). During the second half of 2022, up to 78% of N-IgG positive subjects had knowledge of their infection either by a positive PCR test or at-home antigen test (Figure 1).

## 4. Discussion

Here we show the development of immunity against SARS-CoV-2 in the Finnish adult population from the beginning of the COVID-19 pandemic to the end of 2022. Whilst S-IgG levels increased quickly in 2021 with the introduction of COVID-19 vaccinations, N-IgG seroprevalence remained below 7% until the last quartile of 2021. Age-adjusted N-IgG seroprevalence increased rapidly, and was 31% in Q1/2021 and 54% in Q4/2022, following the waves of infections caused by different Omicron variants^11^. By the end of 2022, S-IgG seroprevalence in the study population was 99%, and 51% had vaccine and infection-induced antibodies, i.e., hybrid immunity.

The N-IgG seroprevalence we observed in 2020 may be an overestimation of all SARS-CoV-2 infections. When applied to low-prevalence populations, even for tests with high, but imperfect specificity, the positive predictive value remains low, i.e. most positive test results are false positives.^12^ On the contrary, later in the pandemic N-IgG seroprevalence may underestimate the number of SARS-CoV-2 infections due to the natural waning of antibody levels^13^. We have previously observed that the sensitivity of the N-IgG assay decreases after six months from infection.^14,15^ By twelve months, N-IgG induced by past SARS-CoV-2 infections are difficult to distinguish from cross-reactive antibodies against seasonal coronaviruses^16,17^ with our assay.^15,18^ We have previously reported that the long-term sensitivity of the S-IgG assay is superior to N-IgG; S-IgG persisted in 97% of subjects at 13 months post-SARS-CoV-2 infection, whilst N-IgG were found in only 36%^18^. For this reason, N-IgG seroprevalence later in the pandemic is not representative of the cumulative number of infections, but rather an insight into more recent ones. Antibodies against SARS-CoV-2 nucleoprotein and spike glycoprotein were initially both used as indicators of previous infection in the serosurvey. However, after COVID-19 vaccinations estimates of infection-induced antibody prevalence could only be based on antibodies against nucleoprotein.

In 2020 most subjects had no antibody-mediated immunity to SARS-CoV-2 and we have previously shown that until June 2020, likely less than 1% of the Finnish population had been infected with SARS-CoV-2^7^. One meta-analysis reported that nucleoprotein antibody seroprevalence increased steadily from 1.4% in March to 4.5% in December 2020 in European high-income countries. These estimates are very similar to the seroprevalences we observed in our study population. In 2021 vaccine-mediated immunity comprised the most of population immunity despite the emergence and dominance of more transmissible Alpha^19,20^ and Delta variants^11,21,22^. We observed that hybrid immunity levels remained low in 2021, indicating few breakthrough infections caused by these variants.

PCR testing capacity was initially limited in Finland and our previous study shows that before April 2020 there were 4–17 infections (95% probability) for every PCR-confirmed infection^7^. The testing capacity then increased and by June 2020 there were 2–3 infections (50% probability) for every PCR-confirmed infection. Although there were some changes in infection testing policy during 2020 and 2021, the finding of this study indicates that the underreporting likely stayed at this modest level until the end of 2021.

In late-2021 PCR testing capacity was surpassed due to the sharp increase in cases caused by the Omicron variant, which was more contagious than previous variants and evaded previous immunity^23^. The limited access to PCR tests resulted in a change in the guidelines^8,9^, and at-home antigen tests became the primary diagnostic method in Finland. PCR tests were primarily offered to only those at risk of a severe infection^9^ and laboratory-confirmed cases no longer represented the total disease burden. For this reason, serosurveillance became the best tool to follow the dynamics of the COVID-19 epidemic in Finland. The switch to the Omicron variant dominance in Finland^11^ was followed by a 6% to 31% jump in N-IgG seroprevalence. Another study conducted in the greater-Helsinki area, Finland, observed very similar (28%) N antibody seroprevalence in March 2022.^24^ In addition, a recent meta-analysis reported an average of 7% N antibody seroprevalence for high-income European countries in December 2021 and a rise to 48% by March 2022^3^, which is slightly higher than what we observed. Furthermore, breakthrough infections caused by the Omicron variants^25,26^ are also apparent in our data; hybrid immunity rates increased from 4% to 31% between Q4/2021 and Q1/2022, and during this time there was a clear shift from only vaccine-induced immunity to hybrid immunity.

One key limitation of this study is that although we use a highly specific and sensitive S-IgG assay^14^ which can detect antibodies for over a year after infection,^15,18^ spike antibodies cannot be used to assess SARS-CoV-2 infection-induced immunity after early 2021 due to mass COVID-19 vaccinations. Estimations of infection-induced antibody seroprevalence can thereafter only be based on N-IgG, which depicts only recent infections. It is also essential to highlight that the estimates of the prevalence of hybrid immunity presented here are based solely on antibody and vaccine registry data and do not take into account cell-mediated immunity. Another key limitation is the overall low participation rate (28%) and therefore possible selection bias, as willingness to participate may be associated with the likelihood of a previous SARS-CoV-2 infection or COVID-19 vaccination. Furthermore, the low participation rate of unvaccinated individuals did not allow us to make reliable estimations of infections for that subpopulation. The participation rate was lower in younger age groups which we controlled for using adjusted analyses, but we were unable to adjust for differences in seroprevalences between regions. Half of our samples were from the capital region, which accounts for less than third of the total population of Finland, and which initially had the highest incidence of confirmed infections. As individuals living in this region are overrepresented, we may overestimate the population immunity. Also, population subgroups who are not native in either of Finland’s two official languages were underrepresented, which may have led to an underestimation of population immunity during the beginning of the epidemic, as COVID-19 incidence among these groups was higher in comparison to the native speakers.

The strengths of this study include the usage of well-validated and specific in-house antibody tests and population surveys spanning the Finnish population. Moreover, access to nationwide high-quality records on PCR-confirmed infections and COVID-19 vaccinations provides valuable information for estimating assay sensitivity and population-wide hybrid immunity. By monitoring the seroprevalence over nearly three years, this study provides a longitudinal view of the COVID-19 epidemic and the evolution of immunity in the Finnish adult population. Previously published serosurveillance reports in Finland have been focused on shorter time periods^7,27^, and seroprevalence in 2021 has not been reported elsewhere, making our study unique. Furthermore, at the time of writing this manuscript, we are among the first to report SARS-CoV-2 seroprevalence in late 2022.

We have combined seroprevalence with data from the vaccination register and the infectious disease register and compared it to the genome sequencing data^11^ of SARS-CoV-2 variants from the same period. This has allowed us to monitor the population immunity as vaccinations have progressed and during the dominance of different SARS-CoV-2 variants in Finland. Our results indicate that the containment measures in combination with high vaccine coverage were able to limit the spread of SARS-CoV-2 until the emergence of the highly transmissible Omicron variants. In 2022 the proportion of subjects with hybrid immunity increased, especially among the younger age groups, where more infections have occurred. Shortly after the start of the first Omicron wave, it was possible to observe the quick shift in the number of infections by serology, because most of the population were N-IgG negative before the wave and we were able to collect samples during the wave. When seroprevalence has risen to over 50 percent, it is more difficult to observe changes in COVID-19 incidence by measuring seroprevalence in a cross-sectional study setting. To conclude, the findings of this study are significant for the understanding of the COVID-19 pandemic, and how different variants, containment measures and vaccinations affected the population. Furthermore, this study can be used to inform and improve strategies for preventing the spread of infectious diseases and mitigating the impacts of future outbreaks.

## 5. Methods

### 5.1 Study design and population

We invited a total of 34 619 subjects by regular mail to participate in the study in forty sequential random population surveys between April 2020 and December 2022. The size of each population survey was adapted to the expected true seroprevalence in the population to achieve a predefined 2–3 percentage point accuracy in the 90% interval estimate of seroprevalence. Due to the properties of the binomial distribution, the required sample size is highest when the true seroprevalence is close to 50%, and lowest when it is close to 0% or 100%. Initially, the target size of each survey was 750, which is sufficient for 2 percentage point accuracy in the estimate in a population where seroprevalence is low (<10%). As seroprevalence estimates remained below 5%, the size of the surveys was lowered (Table S1). As the seroprevalence was expected to have increased by 2022 due to the emergence of more infectious variants, the size of the sample was increased accordingly, with a target size of 1300 per survey. This latter sample size is sufficient for 2 percentage point accuracy when the prevalence is below 20%, and 3 percentage point accuracy when prevalence is 50%. Note that in the current study, we report 95% confidence intervals for seroprevalence.

During each survey, subjects living within the five largest healthcare districts in Finland were randomly selected from the Finnish population registry. During 2020-2021 invitations were sent to 18-70-year-olds and in 2022 to 18-85-year-olds. The Helsinki and Uusimaa healthcare district had the biggest weight in samples (53% of all invitations) due to its larger population and it being the area where the outbreak first started in Finland, subsequently progressing to other areas. Individuals living within institutional care and those previously invited to this study or the follow-up study of the serological population survey of the coronavirus epidemic^18^ were excluded. We asked the participants to provide a blood sample for antibody testing at their local healthcare district laboratory. We retrieved information on COVID-19 vaccinations from Finland’s national vaccination register and PCR-confirmed SARS-CoV-2 infections from the national infectious diseases register. The subjects who participated in 2022 were also asked if they had had a positive at-home antigen test and if yes, when.

### 5.2 SARS-CoV-2 fluorescent multiplex immunoassay

We measured the concentration of serum IgG antibodies to SARS-CoV-2 nucleoprotein (N-IgG) and two spike glycoprotein (S-IgG) antigens with an in-house fluorescent multiplex immunoassay (FMIA) as previously described^14^ with slight modifications detailed in the supplementary material. The IgG SARS-CoV-2 FMIA is an accredited assay at the Expert Microbiology Unit of the Finnish Institute of Health and Welfare, which is a testing laboratory T077 accredited by FINAS Finnish Accreditation Service, accreditation requirement SFS-EN ISO/IEC 17025. The IgG SARS-CoV-2 FMIA assay has been calibrated to the WHO international standard^28^. We present results separately for the proportion of individuals with N-IgG and S-IgG antibodies (N-IgG seroprevalence, S-IgG seroprevalence).

### 5.3 Definitions for infection-induced and vaccine-induced immunity

We assessed the quality of antibody-mediated immunity by combining N-IgG and S-IgG antibody results with COVID-19 vaccination status as indicated in the national vaccination register via linkage by the unique personal identity code and separated subjects into five non-overlapping categories of immunity: no immunity, infection immunity, vaccination immunity, vaccinated without response, hybrid immunity (Table 2). Results were grouped by sample collection date into year quartiles, with samples collected from January to March comprising Q1, April to June Q2, July to September Q3 and October to December Q4.

**Table 2.** Antibody-mediated immunity and criteria for each category.

| Category | Vaccinated | N-IgG positive | S-IgG positive |
| --- | --- | --- | --- |
| No immunity | No | No | No |
| Only infection | No | Yes and/or S-IgG | Yes and/or N-IgG |
| Only vaccine | Yes | No | Yes |
| Vaccinated without response | Yes | Yes or No | No |
| Hybrid immunity | Yes | Yes | Yes |

### 5.4 Statistical methods

We estimated seroprevalence separately for each year quartile both within age groups (18–29, 30–44, 45–64 and 65–85 years) and across age groups, and report point estimates and 95% confidence intervals. For estimation within age groups, we used the Clopper-Pearson method for confidence intervals. For estimation across age groups, we adjusted for differences in the sample population and our target population (those 18–85 years old living in Finland) by weighting each age group’s seroprevalence estimate by the age group’s population proportion. The age-adjusted estimates and their confidence intervals were based on logistic regression and a Wald-type interval estimate constructed on the log-odds scale, as implemented in the survey R package^29^.

### 5.5 Informed consent and human experimentation guidelines

Participation was voluntary. The study protocol was approved by the ethical committee of the Hospital District of Helsinki and Uusimaa and registered under the study protocol HUS/1137/2020. Written Informed consent was obtained from all study subjects before sample collection.

### 5.6 Data availability

The data and code are available from the corresponding author upon reasonable request.

## Data Availability

The data and code are available from the corresponding author upon reasonable request.

## 6. Author contributions

M.M., A.A.P., H.N. and L.L. designed the study. M.M., N.E and A.A.P. supervised the study conduct. A.S. developed the assay and performed laboratory analyses. A.S. and T.N. analysed the data. A.S., T.N., M.M. and N.E wrote the manuscript, and all authors contributed to the text and approved the final version of the manuscript.

## 7. Acknowledgements

We thank Leena Saarinen, Marja Suorsa, Lotta Hagberg, Saara Suopanki, Raisa Hanninen and Anu Haveri for laboratory analyses, Camilla Virta for work and support in assay development, Marja Leinonen, Arja Rytkönen, Hanna Valtonen, Jenni Krogell, Susanna Celik, Janne Salimäki, Oona Liedes, Saimi Vara, Mervi Lasander and Pirjo Tarkiainen for sample and consent handling, Joni Niemi and Anita Nieminen for sample management, Esa Ruokokoski, Dennis Ahlfors, Juha Oksanen, Timo Koskenniemi, Tommi Korhonen and Mikko Aura for data management, study coordinators and investigators Elina Isosaari, Ritva Syrjänen, Heta Nieminen, Päivi Siren and Maila Kyrölä, the Study Steering committee Terhi Kilpi, Jukka Jokinen, Carita Savolainen-Kopra, Tuija Leino, Simopekka Vänskä, Jaana Halonen, Otto Helve and Mia Kontio for participation in the clinical study design, Erika Lindh and Niko Tervo for providing SARS-CoV-2 variant surveillance data, the countless others who participated in sequencing and sample collection all over Finland and the participants of this study.

## 10. Materials and correspondence

The data and code are available from the corresponding author upon reasonable request.

## References

1. Chan, J. F.-W. et al. A familial cluster of pneumonia associated with the 2019 novel coronavirus indicating person-to-person transmission: a study of a family cluster. The Lancet 395, 514–523 (2020).

2. Msemburi, W. et al. The WHO estimates of excess mortality associated with the COVID-19 pandemic. Nature 613, 130–137 (2023).

3. Bergeri, I. et al. Global SARS-CoV-2 seroprevalence from January 2020 to April 2022: A systematic review and meta-analysis of standardized population-based studies. PLOS Med. 19, e1004107 (2022).

4. Wei, Y. et al. Global COVID-19 Pandemic Waves: Limited Lessons Learned Worldwide over the Past Year. Engineering 13, 91–98 (2022).

5. WHO Coronavirus (COVID-19) Dashboard. https://covid19.who.int.

6. Serological population study of the coronavirus epidemic - THL. Finnish Institute for Health and Welfare https://thl.fi/en/web/thlfi-en/research-and-development/research-and-projects/serological-population-study-of-the-coronavirus-epidemic.

7. Nieminen, T. A. et al. Underreporting of SARS-CoV-2 infections during the first wave of the 2020 COVID-19 epidemic in Finland - Bayesian inference based on a series of serological surveys. 2023.02.15.23285941 Preprint at https://doi.org/10.1101/2023.02.15.23285941 (2023).

8. National strategy for COVID-19 testing and contact tracing updated due to changes in epidemiological situation. Ministry of Social Affairs and Health https://stm.fi/en/-/national-strategy-for-covid-19-testing-and-contact-tracing-updated-due-to-changes-in-epidemiological-situation.

9. Strategy for COVID-19 testing and contact tracing updated to prevent spread of disease particularly among risk groups. Ministry of Social Affairs and Health https://stm.fi/-/koronatestaus-ja-jaljitysstrategia-on-paivitetty-tavoitteena-ehkaista-taudin-leviamista-erityisesti-riskiryhmissa?languageId=en_US.

10. Vauhkonen, H. et al. Introduction and Rapid Spread of SARS-CoV-2 Omicron Variant and Dynamics of BA.1 and BA.1.1 Sublineages, Finland, December 2021. Emerg. Infect. Dis. 28, 1229–1232 (2022).

11. SARS-CoV-2-koronaviruksen genomiseuranta - THL. Terveyden ja hyvinvoinnin laitos https://thl.fi/fi/web/infektiotaudit-ja-rokotukset/ajankohtaista/ajankohtaista-koronaviruksesta-covid-19/muuntuneet-koronavirukset/sars-cov-2-koronaviruksen-genomiseuranta.

12. Rogan, W. J. & Gladen, B. Estimating prevalence from the results of a screening test. Am. J. Epidemiol. 107, 71–76 (1978).

13. Fenwick, C. et al. Changes in SARS-CoV-2 Spike versus Nucleoprotein Antibody Responses Impact the Estimates of Infections in Population-Based Seroprevalence Studies. J. Virol. 95, e01828–20 (2021).

14. Solastie, A. et al. A Highly Sensitive and Specific SARS-CoV-2 Spike- and Nucleoprotein-Based Fluorescent Multiplex Immunoassay (FMIA) to Measure IgG, IgA, and IgM Class Antibodies. Microbiol. Spectr. 9, e0113121 (2021).

15. Dub, T. et al. High secondary attack rate and persistence of SARS-CoV-2 antibodies in household transmission study participants, Finland 2020-2021. Front. Med. 9, 876532 (2022).

16. Dobaño, C. et al. Immunogenicity and crossreactivity of antibodies to the nucleocapsid protein of SARS-CoV-2: utility and limitations in seroprevalence and immunity studies. Transl. Res. 232, 60 (2021).

17. Tamminen, K., Salminen, M. & Blazevic, V. Seroprevalence and SARS-CoV-2 cross-reactivity of endemic coronavirus OC43 and 229E antibodies in Finnish children and adults. Clin. Immunol. 229, 108782 (2021).

18. Haveri, A. et al. Persistence of neutralizing antibodies a year after SARS-CoV-2 infection in humans. Eur. J. Immunol. 51, 3202–3213 (2021).

19. Davies, N. G. et al. Estimated transmissibility and impact of SARS-CoV-2 lineage B.1.1.7 in England. Science 372, eabg3055 (2021).

20. Lyngse, F. P. et al. Increased transmissibility of SARS-CoV-2 lineage B.1.1.7 by age and viral load. Nat. Commun. 12, 7251 (2021).

21. Hart, W. S. et al. Generation time of the alpha and delta SARS-CoV-2 variants: an epidemiological analysis. Lancet Infect. Dis. 22, 603–610 (2022).

22. Ong, S. W. X. et al. Clinical and Virological Features of Severe Acute Respiratory Syndrome Coronavirus 2 (SARS-CoV-2) Variants of Concern: A Retrospective Cohort Study Comparing B.1.1.7 (Alpha), B.1.351 (Beta), and B.1.617.2 (Delta). Clin. Infect. Dis. Off. Publ. Infect. Dis. Soc. Am. 75, e1128–e1136 (2022).

23. Carabelli, A. M. et al. SARS-CoV-2 variant biology: immune escape, transmission and fitness. Nat. Rev. Microbiol. 1–16 (2023) doi:10.1038/s41579-022-00841-7.

24. Ahava, M. J. et al. Rapid increase in SARS-CoV-2 seroprevalence during the emergence of Omicron variant, Finland. Eur. J. Clin. Microbiol. Infect. Dis. 1–3 (2022) doi:10.1007/s10096-022-04448-x.

25. Wilhelm, A. et al. Limited neutralisation of the SARS-CoV-2 Omicron subvariants BA.1 and BA.2 by convalescent and vaccine serum and monoclonal antibodies. eBioMedicine 82, 104158 (2022).

26. Pulliam, J. R. C. et al. Increased risk of SARS-CoV-2 reinfection associated with emergence of Omicron in South Africa. Science 376, eabn4947 (2022).

27. Tähtinen, P. A. et al. Low pre-vaccination SARS-CoV-2 seroprevalence in Finnish health care workers: a prospective cohort study. Infect. Dis. Lond. Engl. 54, 448–454 (2022).

28. Mattiuzzo, G. et al. Establishment of the WHO International Standard and Reference Panel for anti-SARS-CoV-2 antibody. World Health Organ. Expert Comm. Biol. Stand. 60 doi:WHO/BS/2020.2403.

29. Lumley, T. Package ‘survey’. Analysis of Complex Survey Samples. https://cran.r-project.org/web/packages/survey/survey.pdf (2021).

