## supplementary material, Table S1, Table S2, Table S3, Table S4, Table S5 for "Changes in SARS-CoV-2 seroprevalence and population immunity in Finland, 2020–2022"

### 1. Supplementary information

#### 1.1 FMIA assay specifications and thresholds for positivity

Assay specificity and sensitivity were determined by analysing sera (n=402) collected in mid-2019 (negative controls) and sera collected in 2020 from subjects (n=87) with PCR-confirmed COVID-19 14-51 days after symptom onset and no COVID-19 vaccination history (positive controls). The positive control sample material has been previously described[1]. Samples collected from April to September 2020 (n=3954) were only analysed for N-IgG, and the samples thereafter were analysed for N-IgG and S-IgG (n=5840).

Samples collected from April 2020 to December 2021 (n=7039) were analysed as duplicates diluted 1:100. The threshold for N-IgG positivity during this period was >6.7 BAU/ml resulting in 100% specificity and 95.8% sensitivity. Samples with >0.9 BAU/ml IgG to receptor binding domain and >1.6 BAU/ml IgG to full-length spike glycoprotein were considered S-IgG positive and the assay specificity and sensitivity were 100%.

Samples collected in 2022 (n=2755) were analysed as duplicates diluted 1:100 and 1:1600. The threshold for N-IgG positivity during this period was >7 BAU/ml resulting in 92.8% specificity and 100% sensitivity. Samples with >3 BAU/ml IgG to receptor binding domain and >2 BAU/ml IgG to full-length spike glycoprotein were considered S-IgG positive, and the assay specificity and sensitivity were 100%.

#### 1.2 Supplementary references

1. Dub, T. *et al.* High secondary attack rate and persistence of SARS-CoV-2 antibodies in household transmission study participants, Finland 2020-2021. *Front. Med.* **9**, 876532 (2022).

#### 1.3 Tables

Table S1. Description of the random population surveys

| Survey | Week/year | Sample size total/per district | Invited subjects | Participated subjects | Participation rate | Healthcare district |
|---|---|---|---|---|---|---|
| 1 | 14/2020 | 750 | 733 | 468 | 64% | HUS |
| 2 | 15/2020 | 750 | 728 | 451 | 62% | HUS |
| 3 | 16/2020 | 1000/200 | 989 | 509 | 51% | All five |
| 4 | 17/2020 | 1000/200 | 980 | 469 | 48% | All five |
| 5 | 17/2020 | 1000/200 | 980 | 480 | 49% | All five |
| 6 | 18/2020 | 750/150 | 738 | 313 | 42% | All five |
| 7 | 19/2020 | 400/80 | 392 | 162 | 41% | All five |
| 8 | 21/2020 | 400/80 | 394 | 145 | 37% | All five |
| 9 | 22/2020 | 400/80 | 398 | 171 | 43% | All five |
| 10 | 22/2020 | 900/500§/80 | 876 | 314 | 36% | All five |
| 11 | 25/2020 | 400/80 | 391 | 132 | 34% | All five |
| 12 | 29/2020 | 400/80 | 391 | 130 | 33% | All five |
| 13 | 31/2020 | 400/80 | 396 | 138 | 35% | All five |
| 14 | 33/2020 | 400* | 392 | 113 | 29% | All five |
| 15 | 35/2020 | 400* | 390 | 113 | 29% | All five |
| 16 | 37/2020 | 400* | 392 | 112 | 29% | All five |
| 17 | 39/2020 | 400* | 387 | 113 | 29% | All five |
| 18 | 41/2020 | 400* | 392 | 107 | 27% | All five |
| 19 | 43/2020 | 400* | 392 | 104 | 27% | All five |
| 20 | 45/2020 | 600*# | 599 | 174 | 29% | All five |
| 21 | 47/2020 | 600*# | 600 | 170 | 28% | All five |
| 22 | 49/2020 | 600*# | 601 | 171 | 28% | All five |
| 23 | 52/2020 | 600*# | 594 | 164 | 28% | All five |
| 24 | 02/2021 | 600*# | 595 | 160 | 27% | All five |
| 25 | 05/2021 | 600*# | 597 | 192 | 32% | All five |
| 26 | 07/2021 | 600*# | 588 | 178 | 30% | All five |
| 27 | 09/2021 | 600*# | 594 | 184 | 31% | All five |
| 28 | 11/2021 | 400 | 382 | 111 | 29% | HUS |
| 29 | 14/2021 | 600*# | 599 | 181 | 30% | All five |
| 30 | 15/2021 | 400 | 391 | 119 | 30% | HUS |
| 31 | 17/2021 | 600*# | 601 | 147 | 24% | All five |
| 32 | 19/2021 | 400 | 390 | 101 | 26% | HUS |
| 33 | 21/2021 | 600*# | 603 | 123 | 20% | All five |
| 34 | 23/2021 | 400 | 385 | 83 | 22% | HUS |
| 35 | 27/2021 | 400 | 386 | 86 | 22% | HUS |
| 36 | 41/2021 | 750 | 728 | 153 | 21% | HUS |
| 37 | 10/2022 | 2500 | 2426 | 377 | 16% | HUS |
| 38 | 17/2022 | 2500 | 2424 | 458 | 19% | HUS |
| 39 | 19/2022 | 4000/1000 | 3929 | 731 | 19% | All excl. HUS |
| 40 | 40/2022 | 6500/2500§/1000 | 6358 | 1190 | 19% | All five |

HUS= Helsinki and Uusimaa healthcare district
*sample size per district adjusted by population
### sample size for HUS district doubled
§ sample size for HUS district

Table S2. The number of participants per year and month.

| | | 2020 | 2021 | 2022 |
|---|---|---|---|---|
| Q1 | January | 0 | 53 | 1 |
| | February | 0 | 413 | 0 |
| | March | 0 | 418 | 168 |
| Q2 | April | 1460 | 314 | 87 |
| | May | 1305 | 319 | 456 |
| | June | 685 | 215 | 791 |
| Q3 | July | 201 | 43 | 38 |
| | August | 244 | 83 | 14 |
| | September | 242 | 3 | 8 |
| Q4 | October | 245 | 13 | 258 |
| | November | 261 | 132 | 879 |
| | December | 382 | 10 | 53 |

Table S3. Study population compared to Uusimaa region and Finland's 18-85-year-old population structure at the end of 2021.

| | | Study population n=9794 | Uusimaa, Finland, 18-85-year-olds (n=1.35 million) | Whole Finland, 18-85-year-olds (n=4.36 million) |
|---|---|---|---|---|
| Age | 18-29 | 11% | 19% | 18 % |
| | 30-44 | 25% | 29% | 25 % |
| | 45-64 | 44% | 32% | 32 % |
| | 65-85 | 19%[a] | 21% | 26 % |
| Sex | Female | 61% | 51% | 50 % |
| | Male | 39% | 49% | 50 % |
| Native language | Finnish or Swedish | 97% | 85% | 91 % |
| | Other | 3% | 15% | 9 % |
| Reside in Uusimaa | | 54% | 100% | 31% |
| Vaccinated for COVID-19[b] | | 96%[d] | 88% | 88% |
| Registered COVID-19 cases[c] | | 2518 (26%)[e] | 457 528 (34%) | 1 219 008 (28%) |

[a] 65-70-year-olds in 2021-2021 and 65-85-year-olds in 2022.
[b] Have received at least one COVID-19 vaccine dose before 15th of December 2021, i.e. would have developed vaccine-mediated immunity by 2022.
[c] January 1st 2020 to December 31st 2022.
[d] Vaccinated before or after sample collection.
[e] Case registered before or after sample collection.

Table S4. The number of serological samples collected in Finland's different healthcare districts per year quartile.

| | | **Number of participants per healthcare district, % of the quartile's samples** | | | | |
|---|---|---|---|---|---|---|
| Year | Quartile | Helsinki and Uusimaa (HUS) | Pirkanmaa | Northern Ostrobothnia | Northern Savonia | Southwest Finland |
| 2020 | Q2, n=3450 | 46% (n=1603) | 15% (n=517) | 12% (n=420) | 14% (n=481) | 12% (n=429) |
| | Q3, n=687 | 35% (n=242) | 20% (n=135) | 16% (n=107) | 13% (n=88) | 17% (n=115) |
| | Q4, n=888 | 65% (n=580) | 13% (n=114) | 7% (n=63) | 7% (n=62) | 8% (n=69) |
| 2021 | Q1, n=884 | 70% (n=618) | 11% (n=101) | 7% (n=62) | 4% (n=37) | 7% (n=66) |
| | Q2, n=847 | 83% (n=705) | 6% (n=50) | 5% (n=44) | 3% (n=22) | 3% (n=26) |
| | Q3, n=129 | 96% (n=124) | 3% (n=4) | 1% (n=1) | 0% (n=0) | 0% (n=0) |
| | Q4, n=155 | 100% (n=155) | 0% (n=0) | 0% (n=0) | 0% (n=0) | 0% (n=0) |
| 2022 | Q1, n=169 | 100% (n=169) | 0% (n=0) | 0% (n=0) | 0% (n=0) | 0% (n=0) |
| | Q2, n=1335 | 48% (n=645) | 13% (n=179) | 11% (n=152) | 14% (n=193) | 12% (n=166) |
| | Q3, n=60 | 33% (n=20) | 20% (n=12) | 5% (n=3) | 15% (n=9) | 27% (n=16) |
| | Q4, n=1190 | 38% (n=451) | 16% (n=191) | 16% (n=188) | 17% (n=202) | 13% (n=158) |
| Total, n=9794 | | 54% (n=5312) | 13% (n=1303) | 11% (n=1040) | 11% (n=1094) | 11% (n=1045) |

Table S5. N-IgG and S-IgG seropositivity by year quartile and age group.

| Year | Quartile | Analysed samples, n | Age | N-IgG positives %, (n) | S-IgG positives %, (n) |
|---|---|---|---|---|---|
| 2020 | Q2 | N-IgG:3450<br>S-IgG:0 | 18-29 | 0.8% (4/523) | NA (0/0) |
| | | | 30-44 | 2.3% (23/1005) | NA (0/0) |
| | | | 45-64 | 2.2% (35/1562) | NA (0/0) |
| | | | 65-70 | 3.1% (11/360) | NA (0/0) |
| | Q3 | N-IgG:687<br>S-IgG:183 | 18-29 | 4.5% (3/66) | 0.0% (0/10) |
| | | | 30-44 | 2.9% (6/204) | 4.8% (3/62) |
| | | | 45-64 | 2.2% (7/317) | 2.3% (2/88) |
| | | | 65-70 | 5.0% (5/100) | 0.0% (0/23) |
| | Q4 | N-IgG:888<br>S-IgG:888 | 18-29 | 6.0% (7/117) | 3.4% (4/117) |
| | | | 30-44 | 2.2% (6/269) | 0.7% (2/269) |
| | | | 45-64 | 2.8% (11/390) | 1.5% (6/390) |
| | | | 65-70 | 0.0% (0/112) | 0.0% (0/112) |
| 2021 | Q1 | N-IgG:884<br>S-IgG:884 | 18-29 | 2.7% (3/112) | 4.5% (5/112) |
| | | | 30-44 | 2.4% (6/249) | 5.2% (13/249) |
| | | | 45-64 | 3.6% (15/419) | 6.9% (29/419) |
| | | | 65-70 | 1.9% (2/104) | 4.8% (5/104) |
| | Q2 | N-IgG:847<br>S-IgG:847 | 18-29 | 4.7% (6/127) | 13.4% (17/127) |
| | | | 30-44 | 7.3% (17/232) | 27.6% (64/232) |
| | | | 45-64 | 4.8% (19/399) | 51.1% (204/399) |
| | | | 65-70 | 3.4% (3/89) | 74.2% (66/89) |
| | Q3 | N-IgG:129<br>S-IgG:129 | 18-29 | 0.0% (0/10) | 70.0% (7/10) |
| | | | 30-44 | 6.3% (3/48) | 77.1% (37/48) |
| | | | 45-64 | 3.6% (2/55) | 90.9% (50/55) |
| | | | 65-70 | 0.0% (0/16) | 93.8% (15/16) |
| | Q4 | N-IgG:155<br>S-IgG:155 | 18-29 | 7.1% (1/14) | 92.9% (13/14) |
| | | | 30-44 | 0.0% (0/41) | 82.9% (34/41) |
| | | | 45-64 | 12.8% (10/78) | 89.7% (70/78) |
| | | | 65-70 | 4.5% (1/22) | 100.0% (22/22) |
| 2022 | Q1 | N-IgG:169<br>S-IgG:169 | 18-29 | 33.3% (3/9) | 100.0% (9/9) |
| | | | 30-44 | 41.2% (14/34) | 100.0% (34/34) |
| | | | 45-64 | 30.4% (21/69) | 100.0% (69/69) |
| | | | 65-85 | 19.3% (11/57) | 100.0% (57/57) |
| | Q2 | N-IgG:1335<br>S-IgG:1335 | 18-29 | 55.4% (41/74) | 95.9% (71/74) |
| | | | 30-44 | 44.9% (96/214) | 95.3% (204/214) |
| | | | 45-64 | 35.6% (195/548) | 98.2% (538/548) |
| | | | 65-85 | 27.9% (139/499) | 99.6% (497/499) |
| | Q3 | N-IgG:60<br>S-IgG:60 | 18-29 | 100.0% (4/4) | 100.0% (4/4) |
| | | | 30-44 | 64.3% (9/14) | 92.9% (13/14) |
| | | | 45-64 | 38.7% (12/31) | 100.0% (31/31) |
| | | | 65-85 | 36.4% (4/11) | 100.0% (11/11) |
| | Q4 | N-IgG:1190<br>S-IgG:1190 | 18-29 | 71.2% (37/52) | 98.1% (51/52) |
| | | | 30-44 | 55.8% (96/172) | 97.7% (168/172) |
| | | | 45-64 | 53.6% (241/450) | 98.9% (445/450) |
| | | | 65-85 | 39.7% (205/516) | 99.2% (512/516) |
